# Development and Validation of Machine Learning Models for Predicting 13 or More Sections in Mohs Micrographic Surgery

**DOI:** 10.64898/2026.07.20.26358484

**Authors:** Yagiz Alp Aksoy, Simon Lee, Gilberto Moreno-Bonilla

## Abstract

**Background:** Cases requiring 13 or more tissue sections in Mohs micrographic surgery (MMS) demand extended operative time, additional resources, and often specialised closure techniques. Pre-operative identification of such cases would improve surgical scheduling, resource allocation, and patient counselling. We aimed to develop and validate a machine learning prediction tool using pre-operative clinical features to identify cases likely to require ≥13 sections.

**Objectives:** To develop and validate machine learning models for predicting which Mohs procedures will require ≥13 sections, using pre-operative clinical features, and to identify key predictive factors.

**Methods:** We analysed 408 consecutive Mohs procedures with 16 pre-operative clinical variables. Thirty machine learning algorithms were evaluated, including ensemble methods (Stacking, Voting), gradient boosting (XGBoost, LightGBM, CatBoost), neural networks (3-7 layers), support vector machines, and traditional classifiers. Model performance was assessed using 5-fold stratified cross-validation and independent test set evaluation. Feature importance was determined using SHAP (SHapley Additive exPlanations) analysis.

**Results:** The stacking ensemble achieved the highest cross-validation AUC of 0.891 (95% CI: 0.849-0.934) and test AUC of 0.884. Tumour area (cm²), calculated using the ellipse formula to approximate clinical tumour morphology, emerged as the strongest predictor (SHAP importance: 0.141), followed by tumour size dimensions (0.086 and 0.068), aggressive histopathology (0.046), and recurrence status (0.035). Wide neural network architectures (5-layer) outperformed deeper configurations (7-layer). The model demonstrated 70.7% high-confidence predictions with uncertainty <15%.

**Conclusions:** Machine learning models using pre-operative clinical features can accurately predict which Mohs procedures will require 13 or more sections. The stacking ensemble approach provides robust predictions suitable for clinical decision support. External validation in multi-centre cohorts with diverse patient populations and practice patterns is warranted to assess model generalisability.

## 1. Introduction

Mohs micrographic surgery (MMS) represents the gold-standard treatment modality for keratinocyte cancer (KC)^1^, particularly basal cell carcinoma (BCC) and squamous cell carcinoma (SCC) in anatomically critical locations^2, 3^. The technique’s superiority lies in its complete margin assessment and tissue-sparing approach, achieving cure rates exceeding 98% for primary tumours^4^.

However, significant variability exists in the number of sections required across cases, with the number of tissue sections required ranging from a single section to over 40 sections in complex cases. Cases requiring 13 or more sections often necessitate extended operative time, additional resources, and specialised closure techniques^5–8^. Pre-operative identification of cases likely to require ≥13 sections would facilitate optimal resource allocation, improve patient counselling, and enable better surgical planning.

Recent advances in machine learning (ML) have demonstrated considerable promise in dermatological applications, including melanoma detection, dermoscopic image analysis, and treatment outcome prediction^9, 10^. However, the application of ML to predicting section count in Mohs surgery remains largely unexplored. Traditional statistical approaches have identified tumour size, recurrence status, and anatomical location as predictive factors^5, 6^, but comprehensive ML-based predictive models utilising multiple pre-operative variables have not been systematically evaluated.

The objectives of this study were to: (1) develop and validate ML models for predicting which Mohs procedures will require ≥13 sections using pre-operative clinical features; (2) compare the performance of diverse ML algorithms including cutting-edge ensemble methods, gradient boosting, and deep neural networks; and (3) identify the most important predictive features using explainable AI techniques.

## 2. Methods

### 2.1 Study Design and Population

This retrospective cohort study analysed 408 consecutive Mohs procedures performed at The Skin Hospital between 2012 and 2017. The Skin Hospital is the largest dedicated Mohs micrographic surgery centre in Australia, operating across two sites in Sydney, NSW (Darlinghurst and Westmead). It serves as the only accredited Mohs surgical training centre in New South Wales and houses the largest number of fellowship-trained Mohs surgeons in the country. The study was approved by the HREC at the University of Sydney (HREC 2017/021). Patient consent was obtained for data collection and analysis.

Inclusion criteria comprised all consecutive patients undergoing MMS for keratinocyte cancer at both sites. Exclusion criteria included other non-melanoma skin cancers and cases with missing outcome data.

### 2.2 Data Collection and Variables

Clinical data were extracted from patient electronic medical records including communication letters, histopathology reports and MMS surgical files. The primary outcome was a binary classification: ≥13 sections versus <13 sections. This threshold was selected based on the Medicare Benefits Scheme (MBS) item numbers and the highest reimbursement cut-off for MMS that appears to be consistently used at a national level, and representing cases requiring substantially greater operative resources, in a country with one of the highest keratinocyte cancer incidence rates globally^1, 11^.

Sixteen pre-operative predictor variables were included: patient demographics (age, sex), tumour characteristics (tumour size largest diameter and its perpendicular widest size, tumour area calculated using the ellipse formula [π × (length/2) × (width/2)], tumour type, recurrence status, aggressive histopathology), anatomical factors (body site (Head and neck, and other), body zone (High, Medium or Low risk zones), laterality, body unit (scalp, nose, ear, temple, forehead, cheek, pretibial, eye, trunk, eyebrow, neck, penis, foot, lip, hand), surgical factors (surgeon experience, and surgical planning evaluated as see-and-do), and clinical history (biopsy status, smoking history). The outcome measure was the final section count, classified into two groups: cases requiring <13 sections versus those requiring ≥13 sections. Post-operative variables including defect size, closure type, and clearance status were excluded from the prediction model to maintain pre-operative clinical utility.

### 2.3 Machine Learning Pipeline

#### 2.3.1 Data Preprocessing

Missing values in continuous variables (tumour size: n=8, 2.0%; smoking status: n=20, 4.9%) were imputed using median imputation. Categorical variables were encoded using label encoding for ordinal variables and one-hot encoding for nominal variables. Continuous features were standardised using z-score normalisation to ensure comparable scales across algorithms.

#### 2.3.2 Model Development

Thirty ML algorithms were systematically evaluated across six categories (Supplementary Table 1).

#### 2.3.3 Model Evaluation

Data were split into training (80%, n=326) and test (20%, n=82) sets using stratified sampling to preserve class distribution. Model performance was assessed using 5-fold stratified cross-validation on the training set, with final evaluation on the held-out test set.

Performance metrics included: area under the receiver operating characteristic curve (AUC-ROC), accuracy, sensitivity, specificity, F1-score, precision, and Brier score for probability calibration.

Confidence intervals were calculated using bootstrap resampling (n=1000 iterations).

#### 2.3.4 Feature Importance and Model Interpretability

Feature importance was evaluated using multiple approaches: (1) inherent feature importance from tree-based models (RF, XGBoost, LightGBM, CatBoost)^12, 13^; (2) SHAP (SHapley Additive exPlanations) values for model-agnostic interpretation^14^. SHAP analysis provides both local (individual prediction) and global (overall model) interpretability by computing the contribution of each feature to predictions.

#### 2.3.5 Uncertainty Quantification

Prediction uncertainty was estimated using a Bayesian-inspired approach with 20 RF models trained with different random seeds. Uncertainty was quantified as the standard deviation of predictions across models. Cases with prediction uncertainty <15% were classified as high-confidence predictions.

### 2.4 Statistical Analysis

Descriptive statistics were computed for all variables. Continuous variables were assessed for normality using Shapiro-Wilk test. Group comparisons utilised Mann-Whitney U test for continuous variables and chi-square test for categorical variables. Effect sizes were calculated using Cohen’s d for continuous variables and Cramér’s V for categorical variables. Statistical significance was set at p<0.05 (two-tailed). All analyses were performed using Python 3.12 with scikit-learn, XGBoost, LightGBM, CatBoost, and SHAP libraries.

### 2.5 Clinical Decision Support Tool Development

To translate the predictive models into clinical practice, we developed an interactive web-based clinical decision support tool. The application was built using a modern technology stack comprising React 18 with TypeScript for the frontend interface, Node.js with Express for the backend server, and Python with Flask for the machine learning API. The prediction engine implements a weighted ensemble of six calibrated binary classifiers (Logistic Regression, Neural Network, AdaBoost, Extra Trees, SVM-RBF, and Random Forest), with weights proportional to their respective ROC-AUC performance.

The tool provides multiple functionalities: (1) binary classification predicting whether a case will require ≥13 sections; (2) continuous section count estimation with 95% confidence intervals using quantile regression; (3) operating room time estimation based on predicted stage count; (4) expected defect size calculation; and (5) Australian Medicare Benefits Schedule (MBS) billing code estimation (items 31000, 31001, 31002). Technical specifications are detailed in the Supplementary Technical Reference.

## 3. Results

### 3.1 Study Population

A total of 408 Mohs procedures were analysed. The mean patient age was 68.5 ± 12.9 years, with 57.8% male patients. The majority of tumours were located in the head and neck region (93.4%) and were basal cell carcinomas (90.0%). The target outcome was well-balanced, with 195 (47.8%) cases required ≥13 sections and 213 (52.2%) required <13 sections. Baseline characteristics stratified by section count outcome are presented in Table 1. The distribution of key variables including outcome, age, section counts, tumour area, body zone, and tumour type are illustrated in Figure 1.

**Figure 1.**
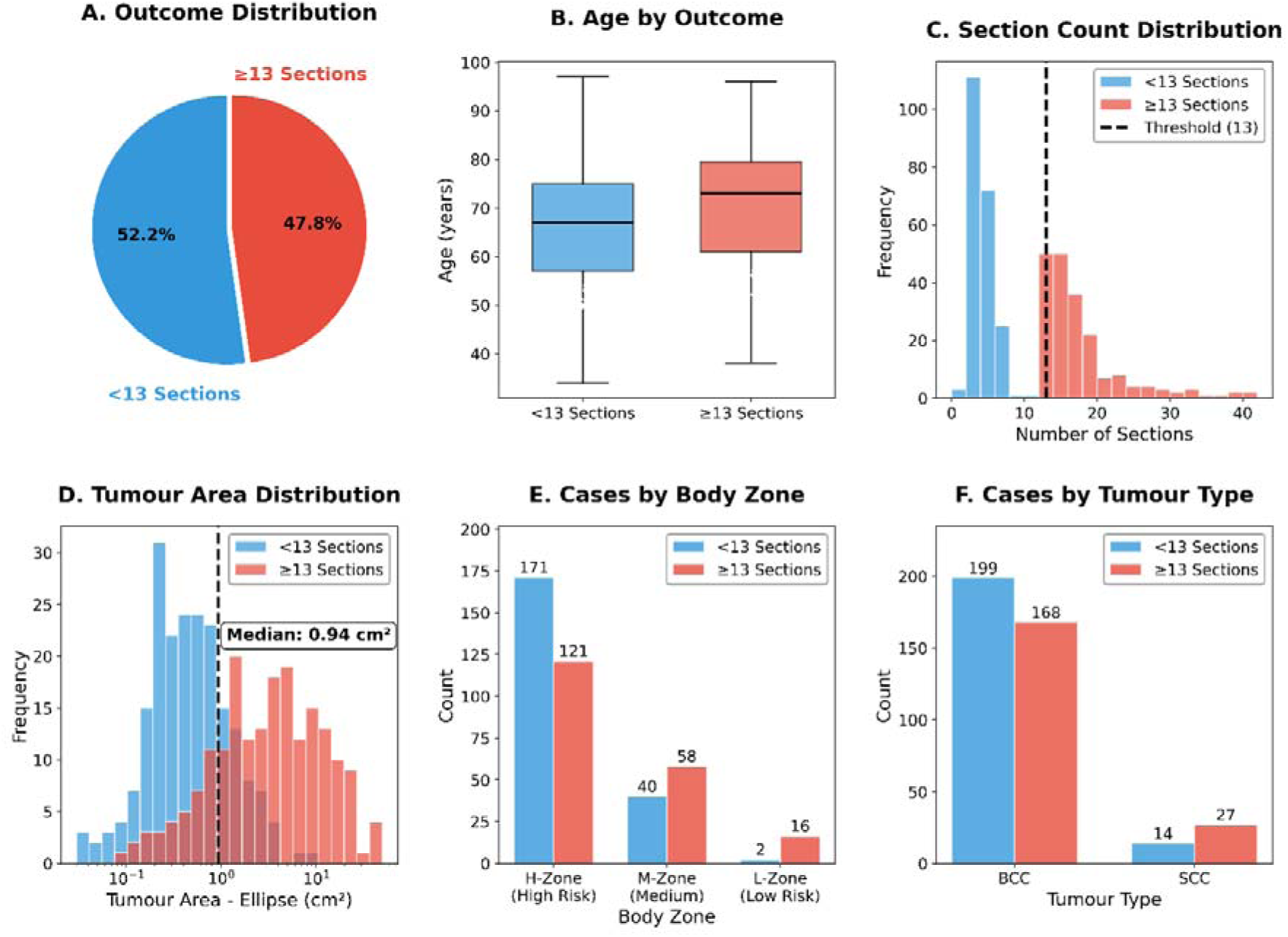
Dataset overview and distribution analysis. (A) Outcome distribution showing 47.8% of cases requiring ≥13 sections (red) and 52.2% requiring <13 sections (blue). (B) Age distribution by outcome group, with mean ages displayed (66.1 years for <13 sections; 71.0 years for ≥13 sections). (C) Section count distribution with threshold line at 13 sections (black dashed line). (D) Tumour area distribution (ellipse formula: π × width/2 × height/2) on logarithmic scale with median of 0.94 cm² indicated. (E) Cases by body zone (H-zone: high-risk, M-zone: medium-risk, L-zone: low-risk) stratified by outcome, with count labels. (F) Cases by tumour type (BCC: basal cell carcinoma, SCC: squamous cell carcinoma) stratified by outcome, with count labels.

**Table 1.**
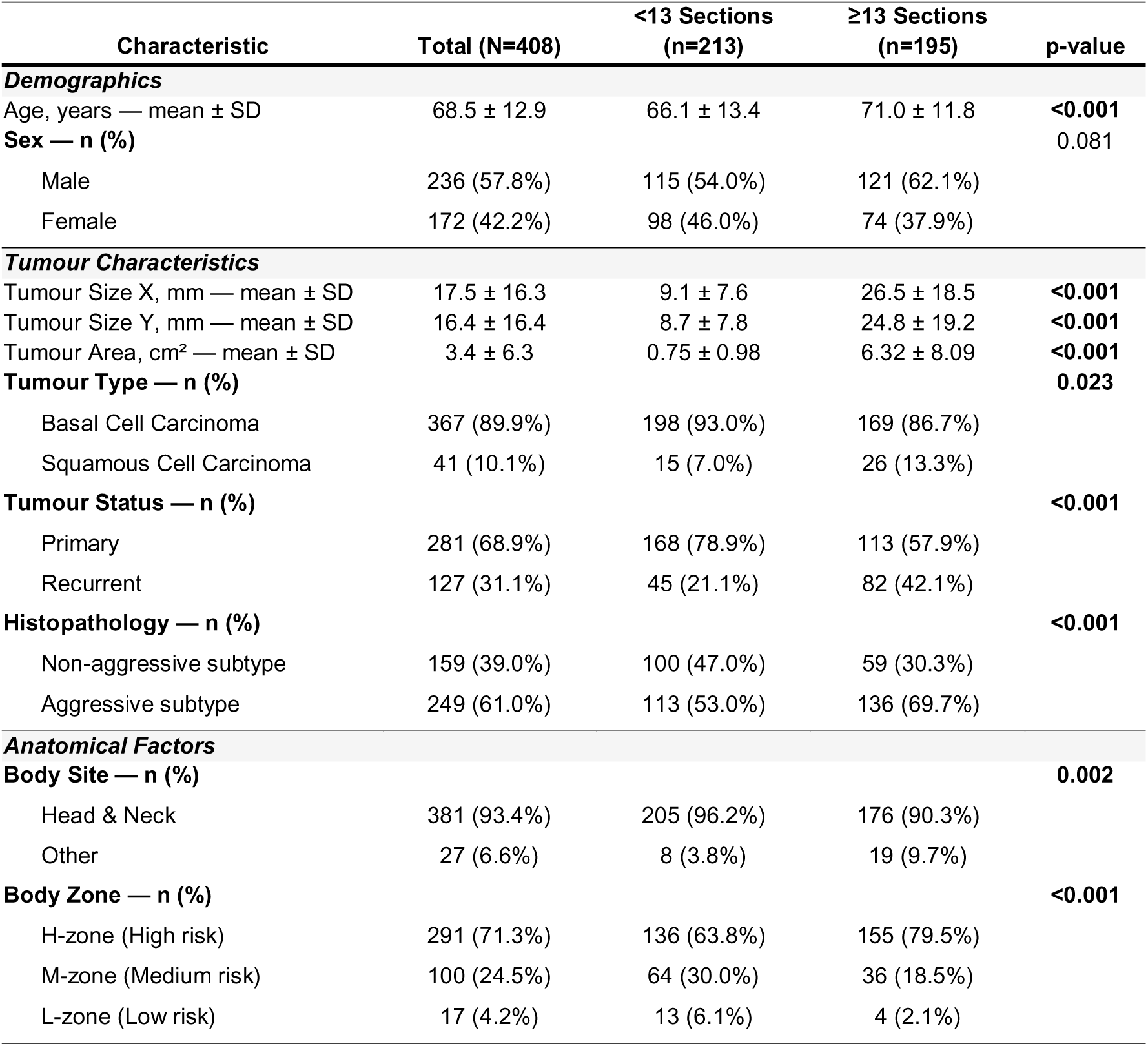
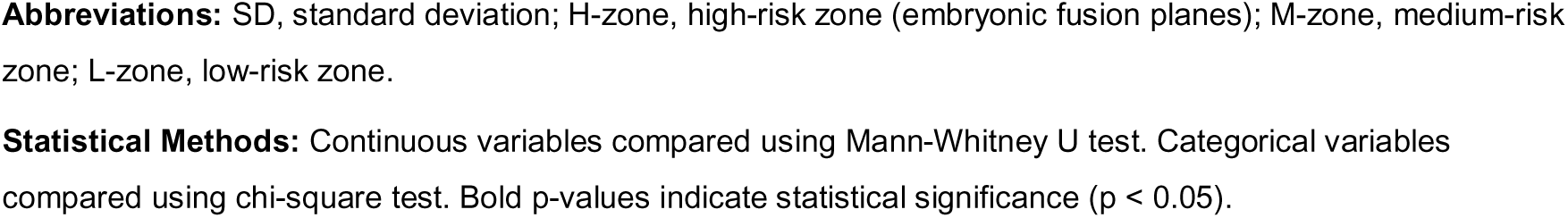
Baseline characteristics of the study population stratified by surgical section count outcome. Continuous variables are presented as mean ± standard deviation or median (interquartile range). Categorical variables are presented as n (%). Statistical comparisons performed using Mann-Whitney U test for continuous variables and chi-square test for categorical variables. BCC, basal cell carcinoma; SCC, squamous cell carcinoma. *p<0.05; **p<0.01; ***p<0.001.

| Characteristic | Total (N=408) | <13 Sections<br>(n=213) | $\geq 13$ Sections<br>(n=195) | p-value |
| --- | --- | --- | --- | --- |
| **Demographics** |  |  |  |  |
| Age, years — mean ± SD | 68.5 ± 12.9 | 66.1 ± 13.4 | 71.0 ± 11.8 | <0.001 |
| Sex — n (%) |  |  |  | 0.081 |
| Male | 236 (57.8%) | 115 (54.0%) | 121 (62.1%) |  |
| Female | 172 (42.2%) | 98 (46.0%) | 74 (37.9%) |  |
| **Tumour Characteristics** |  |  |  |  |
| Tumour Size X, mm — mean ± SD | 17.5 ± 16.3 | 9.1 ± 7.6 | 26.5 ± 18.5 | <0.001 |
| Tumour Size Y, mm — mean ± SD | 16.4 ± 16.4 | 8.7 ± 7.8 | 24.8 ± 19.2 | <0.001 |
| Tumour Area, cm<sup>2</sup> — mean ± SD | 3.4 ± 6.3 | 0.75 ± 0.98 | 6.32 ± 8.09 | <0.001 |
| Tumour Type — n (%) |  |  |  | 0.023 |
| Basal Cell Carcinoma | 367 (89.9%) | 198 (93.0%) | 169 (86.7%) |  |
| Squamous Cell Carcinoma | 41 (10.1%) | 15 (7.0%) | 26 (13.3%) |  |
| Tumour Status — n (%) |  |  |  | <0.001 |
| Primary | 281 (68.9%) | 168 (78.9%) | 113 (57.9%) |  |
| Recurrent | 127 (31.1%) | 45 (21.1%) | 82 (42.1%) |  |
| Histopathology — n (%) |  |  |  | <0.001 |
| Non-aggressive subtype | 159 (39.0%) | 100 (47.0%) | 59 (30.3%) |  |
| Aggressive subtype | 249 (61.0%) | 113 (53.0%) | 136 (69.7%) |  |
| **Anatomical Factors** |  |  |  |  |
| Body Site — n (%) |  |  |  | 0.002 |
| Head & Neck | 381 (93.4%) | 205 (96.2%) | 176 (90.3%) |  |
| Other | 27 (6.6%) | 8 (3.8%) | 19 (9.7%) |  |
| Body Zone — n (%) |  |  |  | <0.001 |
| H-zone (High risk) | 291 (71.3%) | 136 (63.8%) | 155 (79.5%) |  |
| M-zone (Medium risk) | 100 (24.5%) | 64 (30.0%) | 36 (18.5%) |  |
| L-zone (Low risk) | 17 (4.2%) | 13 (6.1%) | 4 (2.1%) |  |

**Abbreviations:** SD, standard deviation; H-zone, high-risk zone (embryonic fusion planes); M-zone, medium-risk zone; L-zone, low-risk zone.
**Statistical Methods:** Continuous variables compared using Mann-Whitney U test. Categorical variables compared using chi-square test. Bold p-values indicate statistical significance ( $p < 0.05$ ).

### 3.2 Statistical Analysis of Clinical Variables

#### Continuous variables

Cases requiring ≥13 sections demonstrated significantly larger tumour areas (6.32 ± 8.09 cm² vs 0.75 ± 0.98 cm², p<0.001, Cohen’s d=0.98). Mean age was higher in the ≥13 sections group (71.0 ± 11.8 years vs 66.1 ± 13.4 years, p<0.001, Cohen’s d=0.39). This age effect was primarily observed within BCC cases (71.0 vs 65.9 years, p<0.001), while SCC cases showed no significant age difference by outcome (70.9 vs 69.2 years, p=0.86). SCC was independently associated with higher rates of ≥13 sections (65.9% vs 45.8% for BCC, p=0.023), though the small SCC sample (n=41) limits subgroup interpretation. Tumour size in both X and Y dimensions was significantly greater in cases requiring ≥13 sections (p<0.001 for both). Univariate statistical analysis results are summarised in Table 2.

**Table 2.**
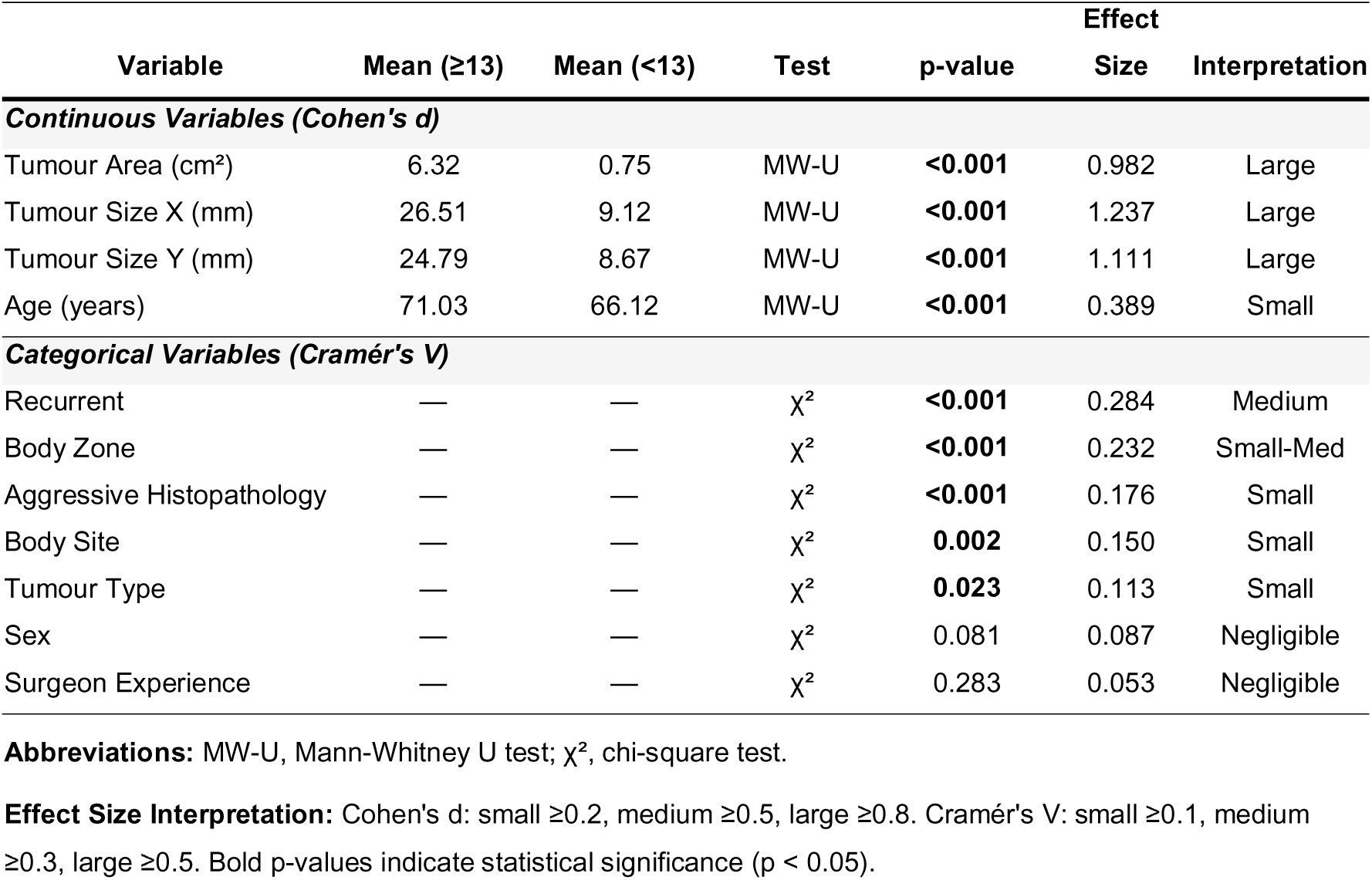
Univariate statistical analysis of predictor variables. Continuous variables analysed using Mann-Whitney U test with Cohen’s d effect size. Categorical variables analysed using chi-square test with Cramér’s V effect size. MW, Mann-Whitney; SD, standard deviation.

#### Categorical variables

Recurrence was significantly associated with requiring ≥13 sections (χ²=32.9, p<0.001, Cramér’s V=0.28). Body zone classification showed significant association (χ²=22.0, p<0.001, Cramér’s V=0.23), with H-zone tumours less likely to require ≥13 sections (42%) compared to M-zone (59%) and L-zone (89%). Aggressive histopathology was associated with requiring ≥13 sections (χ²=12.6, p<0.001, Cramér’s V=0.18). Surgeon experience and see-and-do approach were not significantly associated with outcome (p>0.05).

### 3.3 Machine Learning Model Performance

Thirty ML models were evaluated using 5-fold stratified cross-validation. The complete performance metrics are presented in Table 3.

**Table 3.**
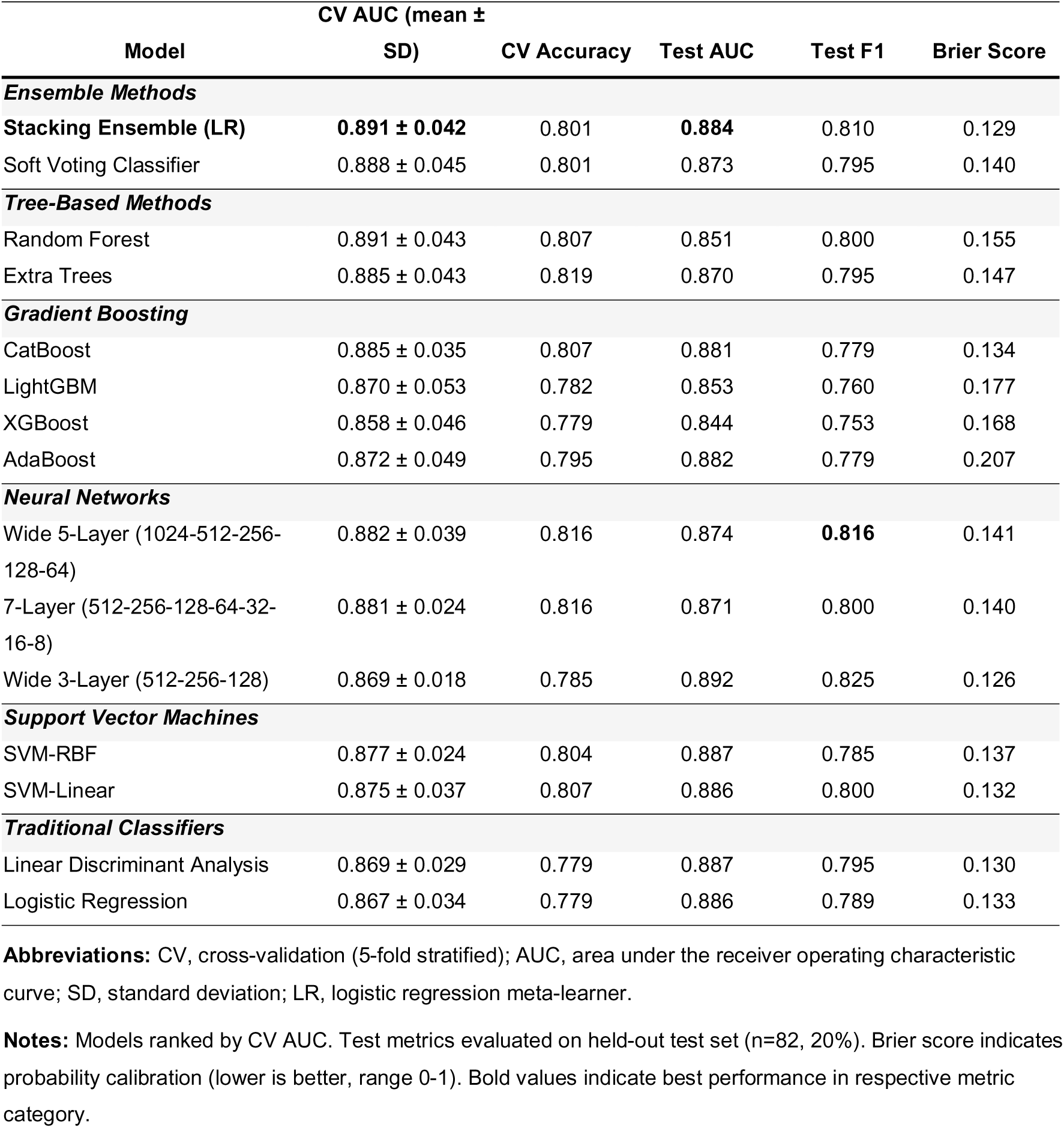
Machine learning model performance comparison. Models ranked by cross-validation (CV) AUC. CV metrics represent mean ± standard deviation across 5 folds. Test metrics evaluated on held-out test set (n=82). AUC, area under the receiver operating characteristic curve; Acc, accuracy; Brier, Brier score (lower is better). Top 20 models shown.

#### Top-performing models

The stacking ensemble with logistic regression meta-learner achieved the highest cross-validation AUC of 0.891 (SD=0.042), with test AUC of 0.884, accuracy of 81.7%, and F1-score of 0.81. Random forest performed comparably (CV AUC=0.891, SD=0.043) but showed slightly lower test performance (test AUC=0.851). CatBoost demonstrated excellent generalisation (CV AUC=0.885, test AUC=0.881) with the best probability calibration (Brier score=0.134). Model performance comparisons are visualised in Figure 2.

**Figure 2.**
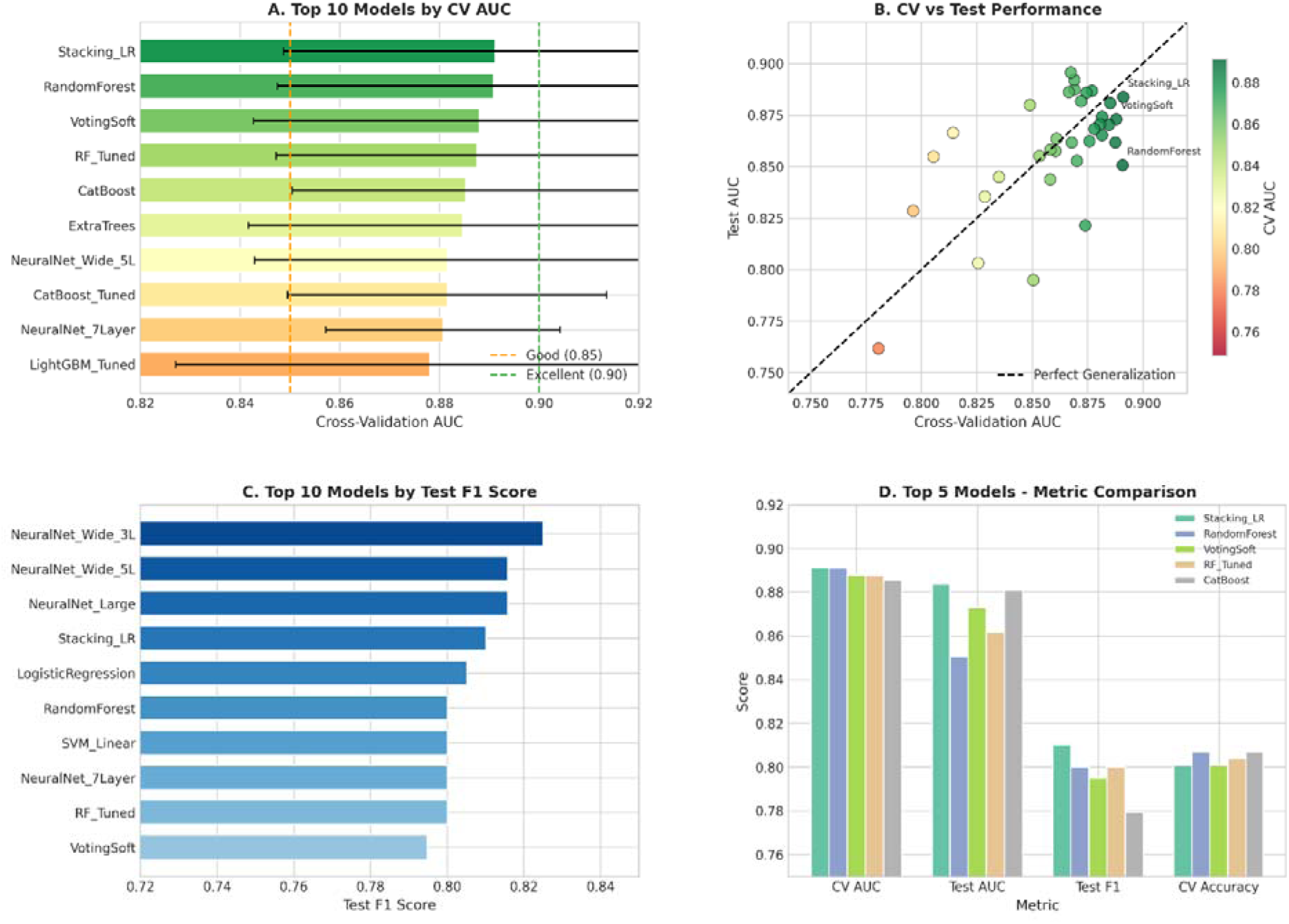
Model performance comparison. (A) Top 10 models ranked by cross-validation AUC with error bars representing standard deviation. Dashed lines indicate good (0.85) and excellent (0.90) performance thresholds. (B) Cross-validation AUC versus test AUC scatter plot demonstrating model generalisation. Points on the diagonal indicate perfect generalisation. (C) Test F1-score comparison. (D) Normalised performance metrics for top 5 models.

#### Neural network architectures

Wide neural networks outperformed deeper configurations. The wide 5-layer architecture (1024-512-256-128-64) achieved CV AUC of 0.882 and test AUC of 0.874, surpassing the 7-layer architecture (CV AUC=0.881, test AUC=0.871). The wide 3-layer network achieved the highest test AUC among neural networks (0.892).

#### Gradient boosting comparison

CatBoost (CV AUC=0.885) slightly outperformed XGBoost (CV AUC=0.858) and LightGBM (CV AUC=0.870) in cross-validation. All three showed strong generalisation to the test set. Receiver operating characteristic and precision-recall curves for top-performing models are shown in Figure 3.

**Figure 3.**
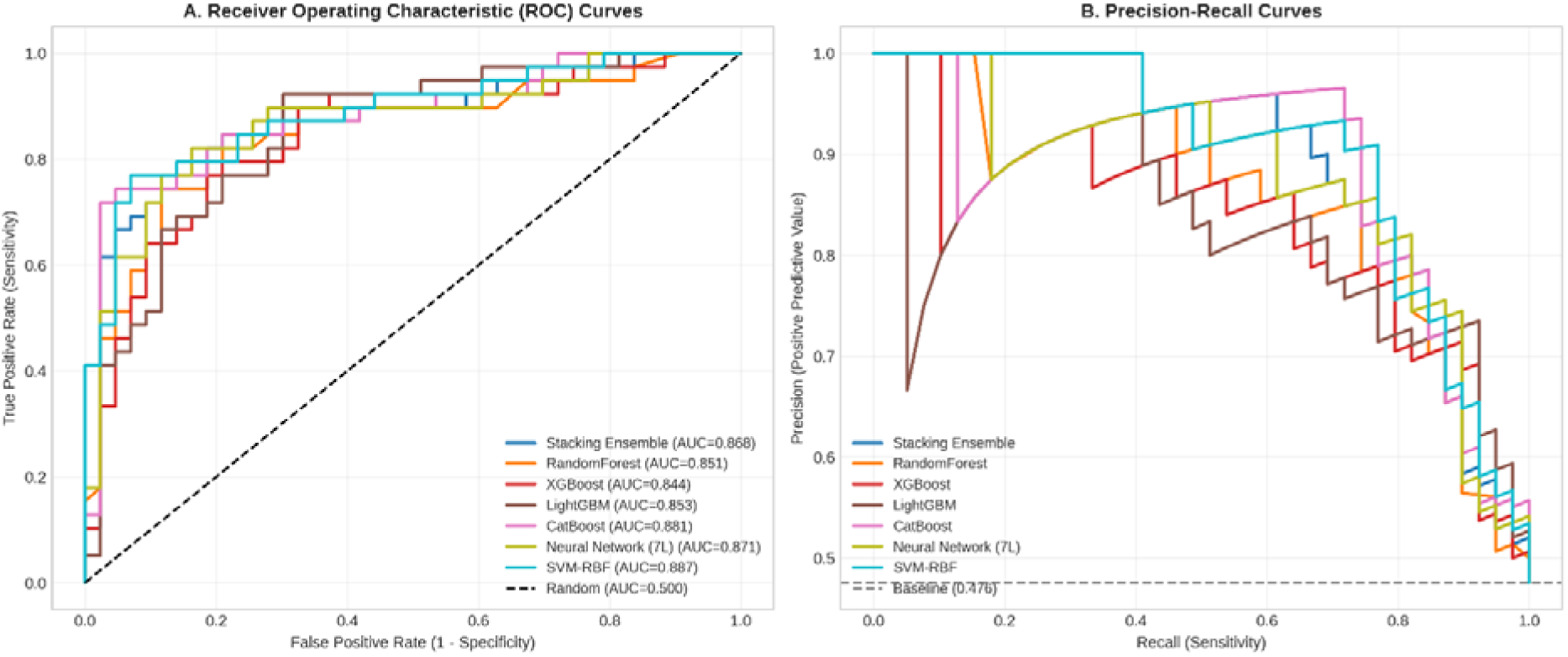
Receiver operating characteristic (ROC) and precision-recall curves for top-performing models. (A) ROC curves showing discrimination ability with AUC values in legend. (B) Precision-recall curves with baseline (prevalence) indicated by dashed line.

### 3.4 Feature Importance Analysis

SHAP analysis showed tumour area as the dominant predictor across all models (mean |SHAP|=0.141). Tumour size X was the second most important feature (0.086), followed by tumour size Y (0.068), aggressive histopathology (0.046), and recurrence status (0.035). Body zone showed moderate importance (0.006), while surgeon experience and see-and-do approach had minimal contribution. Age showed moderate importance (0.035), while surgeon experience and see-and-do approach had minimal contribution (0.008 and 0.005, respectively). Feature importance analysis using multiple methodologies is presented in Figure 4.

**Figure 4.**
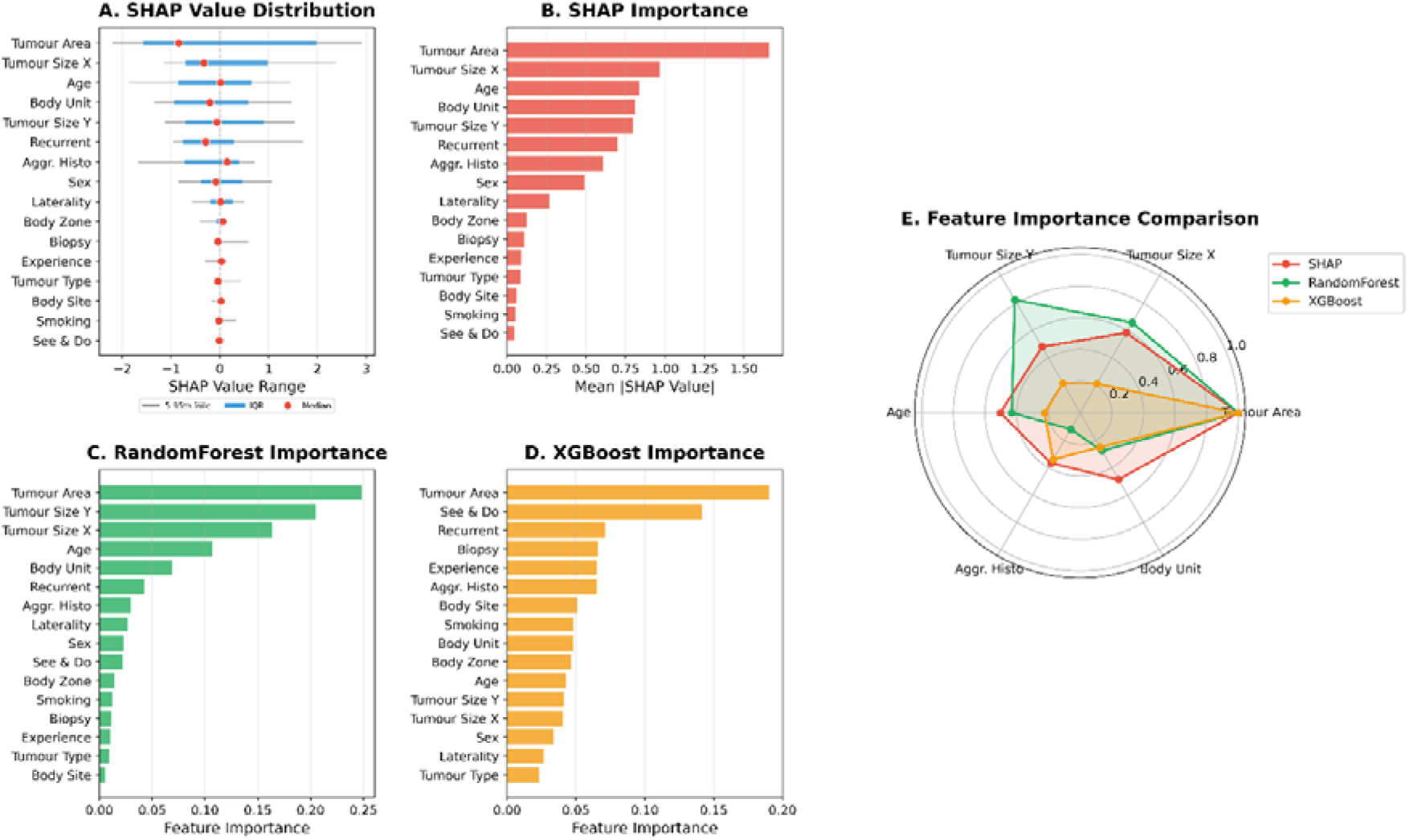
Feature importance analysis using multiple methodologies. (A) SHAP value range by feature showing the distribution of model impact. Horizontal lines represent the 5th-95th percentile range (grey) and interquartile range (blue), with median values indicated by red dots. Features are ordered by importance from bottom to top. (B) SHAP importance showing mean absolute SHAP values for each feature (red bars). (C) RandomForest feature importance rankings (green bars). (D) XGBoost feature importance rankings (orange bars). (E) Radar chart comparing normalised feature importance across the three methods (SHAP, RandomForest, XGBoost) for the top six predictive features. Colours correspond to the respective bar chart colours: red for SHAP, green for RandomForest, and orange for XGBoost. Tumour area consistently emerges as the dominant predictor across all methods.

Feature importance rankings were consistent across RF, XGBoost, LightGBM, and CatBoost models (Spearman correlation ρ>0.85 between all pairs). SHAP dependence plots showed non-linear relationships: tumour area showed a threshold effect at approximately 1.5 cm² (ellipse area), beyond which section count probability increased steeply. SHAP dependence plots demonstrating these relationships are shown in Figure 5.

**Figure 5.**
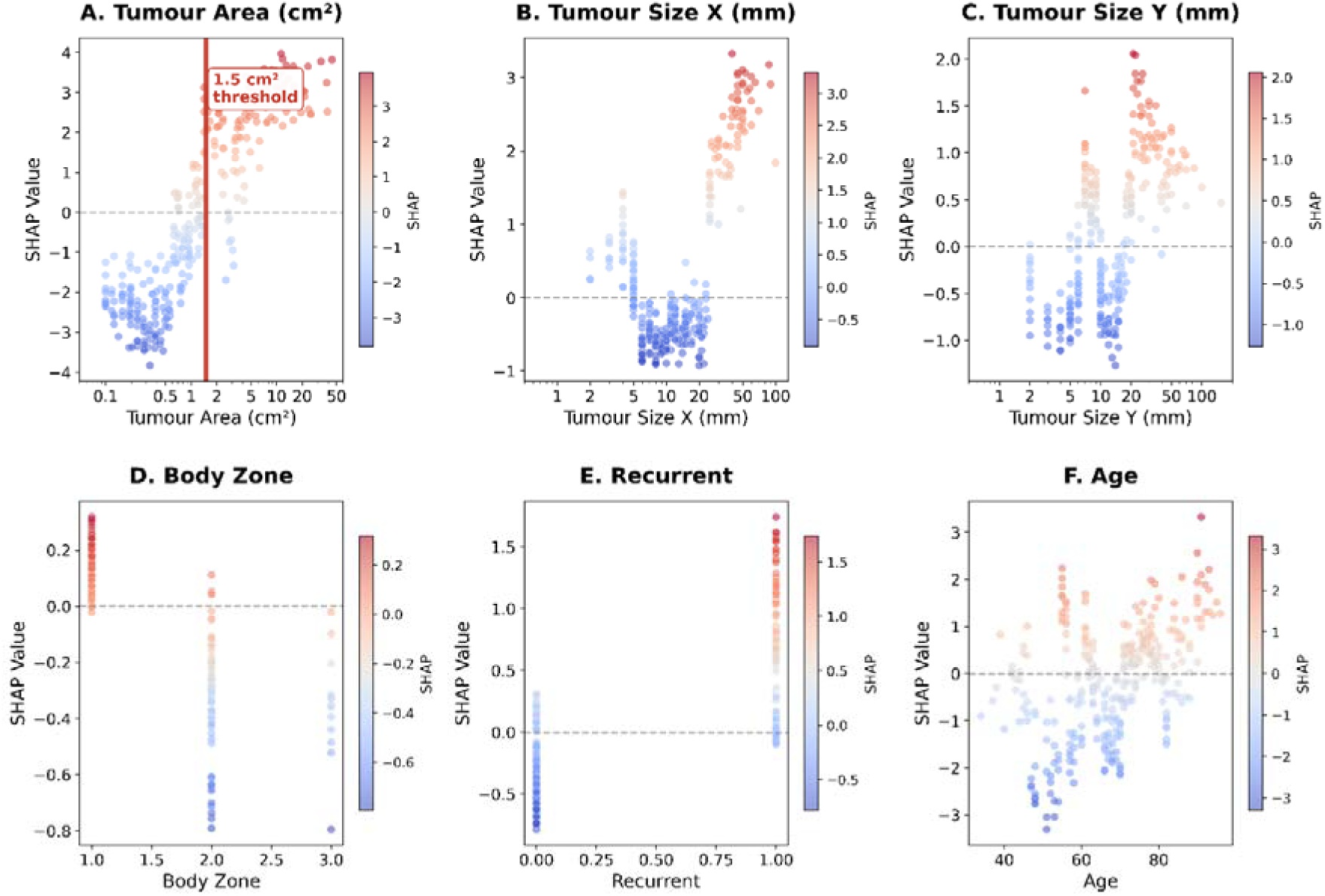
SHAP dependence plots for top predictive features. Each subplot shows the relationship between feature value (x-axis) and SHAP value (y-axis), with point colour indicating the SHAP contribution (red = positive impact towards ≥13 sections; blue = negative impact). Horizontal dashed lines indicate zero SHAP value. (A) Tumour area (cm²) on logarithmic scale with clinical threshold at 1.5 cm² indicated by vertical red line. Values above this threshold show predominantly positive SHAP values, indicating increased likelihood of ≥13 sections. (B) Tumour size X (mm) on logarithmic scale, demonstrating positive correlation with section count probability. (C) Tumour size Y (mm) on logarithmic scale, showing similar positive relationship to tumour dimensions. (D) Body zone (1=H-zone, 2=M-zone, 3=L-zone), showing variable SHAP contributions across anatomical risk zones. (E) Recurrence status (0=primary, 1=recurrent), demonstrating positive SHAP values for recurrent tumours. (F) Age (years), showing modest positive correlation between patient age and section count probability.

**Figure 6.**
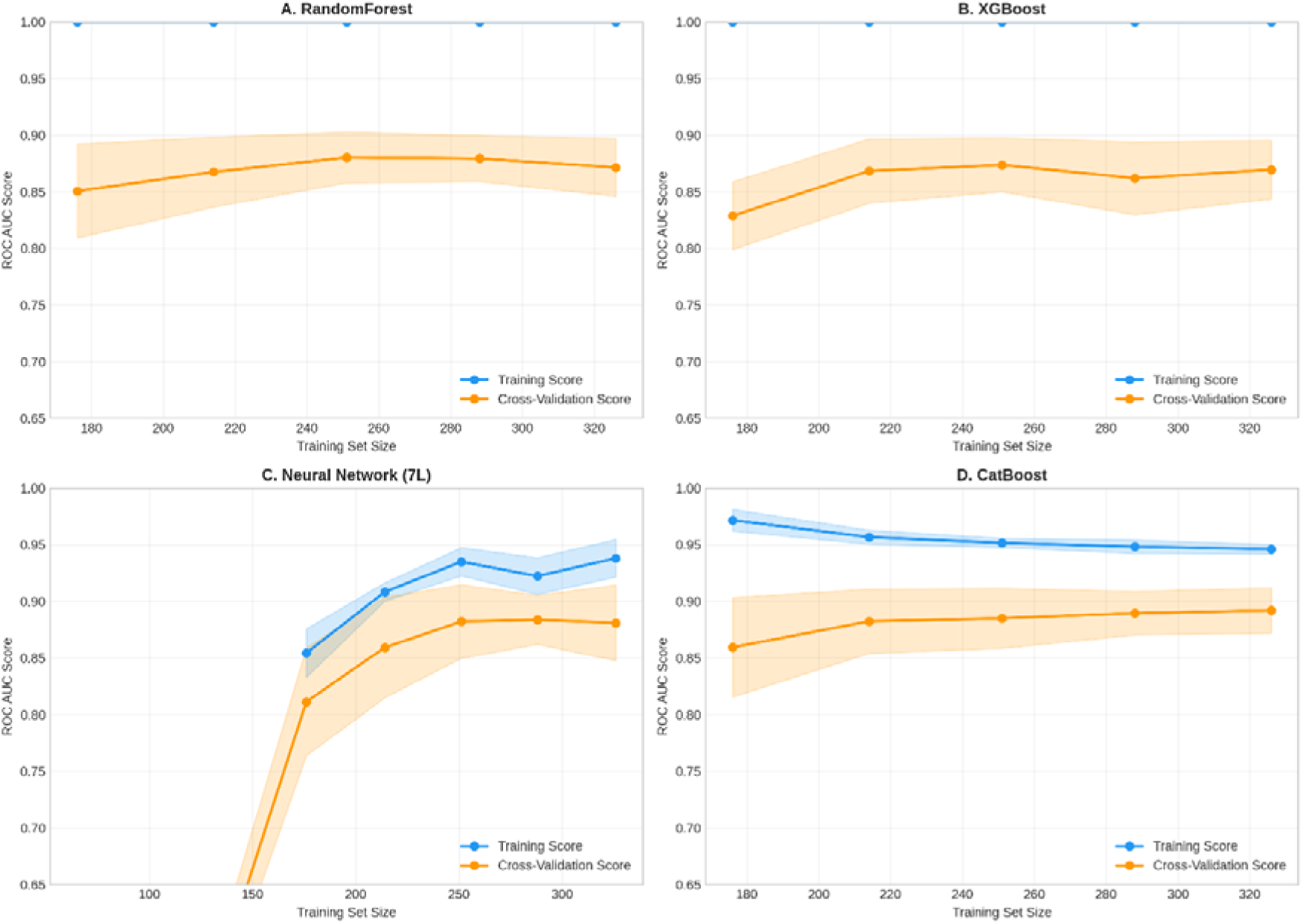
Learning curves for top-performing models. Training set size (x-axis) versus ROC AUC (y-axis). Blue lines represent training scores; orange lines represent cross-validation scores. Shaded regions indicate standard deviation across folds. (A) RandomForest, (B) XGBoost, (C) Neural Network (7-layer), (D) CatBoost.

### 3.5 Model Calibration and Uncertainty

Probability calibration analysis revealed that CatBoost and the stacking ensemble showed optimal calibration, with predicted probabilities closely matching observed frequencies. RF showed slight overconfidence in predictions near 0.5.

Uncertainty quantification demonstrated that 58 of 82 test cases (70.7%) achieved high-confidence predictions (uncertainty <15%). Among high-confidence predictions, accuracy was 91.4%, compared to 70.8% for lower-confidence predictions. Mean prediction uncertainty was 0.121 (range: 0.029-0.246). Calibration curves and uncertainty analysis are presented in Supplementary Figure 1.

### 3.6 Learning Curves and Model Capacity

Learning curve analysis demonstrated that RF and CatBoost reached performance plateaus at approximately 260 training samples, suggesting adequate sample size. Neural networks showed greater variance at smaller sample sizes but achieved comparable performance with full training data.

XGBoost demonstrated slight overfitting tendency, with a larger gap between training and validation scores. Learning curves for top-performing models are shown in Figure 7.

### 3.7 Clinical Decision Support Tool

The trained machine learning models were deployed as an interactive web application, accessible at https://mohs.panacea-i.com. The tool allows clinicians to input pre-operative patient and tumour characteristics and receive real-time predictions including: probability of requiring ≥13 sections, estimated section count with 95% confidence interval, predicted number of stages, estimated operating room time, and expected MBS billing category. The interface displays prediction confidence and provides uncertainty quantification to flag cases requiring additional clinical judgement.

## 4. Discussion

This study presents the first comprehensive evaluation of machine learning approaches for predicting which Mohs procedures will require ≥13 sections. Our findings demonstrate that pre-operative clinical features can achieve high discriminative ability (AUC=0.891) for identifying cases likely to require 13 or more sections, with important implications for clinical practice and resource planning.

### 4.1 Model Performance and Algorithm Selection

The superior performance of ensemble methods aligns with the medical ML literature, where model aggregation consistently outperforms individual algorithms^15, 16^. The stacking ensemble’s advantage likely stems from its ability to leverage complementary strengths of diverse base learners: tree-based models capture non-linear interactions, gradient boosting optimises residual errors, and neural networks model complex feature combinations.

An interesting finding was that wide neural network architectures (5-layer) outperformed deeper configurations (7-layer). This contrasts with computer vision applications where depth correlates with performance, but aligns with recent work on tabular data showing that depth beyond 3-5 layers often provides diminishing returns^17, 18^. Clinical features lack the hierarchical structure of images, where successive layers extract increasingly abstract representations. Wide architectures may better capture feature interactions in tabular medical data.

### 4.2 Clinical Predictors of Section Count

Tumour area emerged as the dominant predictor, consistent with the pathophysiological principle that larger tumours require more extensive excision. Importantly, ≥13 sections serves as a surrogate primarily for tumour size rather than anatomical or surgical complexity per se. The SHAP dependence analysis revealed a threshold effect at approximately 1.5 cm² (ellipse area), suggesting this could serve as a clinical decision point for anticipating procedures requiring 13 or more sections..

Tumour size dimensions and aggressive histopathology were the next most important predictors, while recurrence status also contributed meaningfully. The importance of recurrence reflects the biological behaviour of recurrent tumours, which often exhibit ill-defined margins and subclinical extension due to scarring from prior treatment^19^.

Body zone classification (H-zone, M-zone, L-zone) showed a significant but counterintuitive association with section count. H-zone tumours demonstrated fewer sections (9.0 vs 15.7 in L-zone) and lower rates of ≥13 sections (42% vs 89%), despite being considered anatomically high-risk for recurrence. This apparent paradox is explained by the substantial difference in tumour size between zones: L-zone tumours were approximately six times larger than H-zone tumours (12.55 cm² vs 2.12 cm², p<0.001).

Notably, stages did not differ significantly across zones (p=0.11), while sections-per-stage was markedly lower in H-zone (3.0 vs 8.6, p<0.001). This pattern supports the hypothesis that Mohs surgeons intuitively adopt a more tissue-sparing approach in cosmetically sensitive areas, taking smaller, more conservative cuts per stage. However, the small number of L-zone cases (n=18) limits the strength of conclusions regarding this zone. The higher proportion of H-zone cases (70%) reflects the appropriate utilisation of MMS for tumours in the head and neck region.

The minimal contribution of surgeon experience to the model warrants consideration. This finding may reflect selection bias, where junior Mohs specialists recommending treatment are more comfortable with cases requiring more sections, particularly if reconstruction is conducted by another specialty, thereby confounding the relationship. It has been described that collaboration with other specialties can be driven particularly by anatomic site or by patient’s preference^20, 21^. Future studies examining surgeons’ case selection patterns could provide further information on this relationship.

### 4.3 Clinical Implications

The integration of machine learning models into clinical workflows represents a paradigm shift in surgical planning and resource allocation^22, 23^. Recent advances in medical artificial intelligence have demonstrated significant potential across diverse clinical applications^22^, with high-performance medicine increasingly incorporating predictive analytics^23^. Such tools could facilitate improved patient counselling, enable more accurate operative time estimates, and optimize theatre scheduling.

Successful integration of clinical decision support systems requires careful attention to workflow integration, user training, and ongoing validation^24^.

The demonstrated predictive accuracy of this tool supports several clinical applications:

#### Resource allocation

Pre-operative section count prediction enables scheduling optimisation, with cases likely to require ≥13 sections allocated to longer surgical slots and appropriate staffing levels.

#### Patient counselling

Evidence-based estimates of surgical extent can improve informed consent discussions, particularly for patients with cases with high probability of requiring ≥13 sections.

#### Referral decision-making

Primary care and general dermatology practitioners can utilise predictive tools to identify patients who may benefit from MMS at specialised centres and counsel those who may require a multidisciplinary approach.

The uncertainty quantification component is particularly valuable clinically. Cases with high-confidence predictions (70.7% of the cohort) achieved 91.4% accuracy, providing reliable pre-operative estimates. Lower-confidence predictions flag cases requiring additional clinical judgement.

#### Clinical decision support deployment

To facilitate clinical implementation, we developed and deployed an interactive web-based prediction tool (https://mohs.panacea-i.com). This platform enables clinicians to input pre-operative variables and receive real-time predictions with uncertainty quantification. Beyond binary classification, the tool provides continuous section count estimates with 95% confidence intervals, operating room time predictions, and Australian MBS billing code estimation. Such dynamic tools offer several advantages over static prediction models: they enable real-time decision support at the point of care, automatically incorporate uncertainty quantification to flag borderline cases, and can be readily updated as new data become available or as the model is externally validated. The web-based deployment also facilitates accessibility across devices without requiring software installation, lowering barriers to clinical adoption.

### 4.4 Comparison with Existing Literature

Prior studies have identified tumour size and recurrence as predictive factors using logistic regression, reporting AUC values of 0.70-0.78^7, 8^. Our ML approach demonstrates substantial improvement (AUC=0.891), likely attributable to: (1) utilisation of more comprehensive feature sets; (2) capacity of ML algorithms to capture non-linear relationships and interactions; and (3) ensemble methods combining multiple learning paradigms. Moreno-Bonilla et al.^5^ recently reported that largest tumour diameter, mixed histology, tumour recurrence and age were significant predictors for 13 or more MMS sections using multivariate logistic regression, findings that our ML models corroborate and extend The finding that pre-operative factors explain substantial variance in section count complements literature suggesting that intra-operative factors (margin status, tissue processing) account for residual variability. Future multi-phase prediction approaches incorporating intra-operative data could further improve accuracy.

### 4.5 Strengths and Limitations

#### Strengths

This study provides the most comprehensive ML evaluation for predicting ≥13 sections in Mohs surgery to date, systematically comparing 30 algorithms across multiple categories. The use of SHAP analysis provides interpretable insights essential for clinical adoption. Rigorous validation using cross-validation and held-out test sets reduces overfitting concerns.

#### Limitations

Several limitations warrant consideration. First, while data were derived from a single institution, The Skin Hospital represents the largest Mohs centre in Australia with the highest case volume and largest concentration of fellowship-trained Mohs surgeons nationally, enhancing the generalisability of findings to Australian practice. Nevertheless, external validation in independent cohorts, including international centres, is essential before broader clinical implementation. Second, the threshold of 13 sections, while clinically meaningful, is somewhat arbitrary; alternative thresholds may be appropriate for different institutional contexts. Third, the sample size (n=408), while adequate for ML model development, limits subgroup analyses. Fourth, temporal validation was not performed, and model performance over time requires monitoring. Finally, the exclusion of imaging features (dermoscopy, histopathology images) may limit predictive ceiling; multimodal approaches warrant future investigation.

### 4.6 Future Directions

Future research should prioritise: (1) multi-centre external validation; (2) prospective clinical trials evaluating impact on surgical outcomes and resource utilisation; (3) integration of imaging data using deep learning; (4) prospective evaluation of the deployed clinical decision support tool (https://mohs.panacea-i.com) in clinical practice; (5) investigation of causal inference methods to address selection bias in surgeon assignment; and (6) integration of the web-based prediction tool with electronic medical record systems to enable seamless workflow integration.

## 5. Conclusions

Machine learning models using pre-operative clinical features can accurately predict which Mohs procedures will require 13 or more sections, achieving AUC of 0.867-0.891 with ensemble approaches. Tumour area, tumour size dimensions, and aggressive histopathology are the most important predictive features. Notably, ≥13 sections serves primarily as a surrogate for tumour size rather than surgical complexity per se. Wide neural network architectures outperform deeper configurations for tabular medical data. The demonstrated predictive accuracy and interpretability support clinical implementation for resource planning and patient counselling, pending external validation. A web-based clinical decision support tool implementing these models is freely available at https://mohs.panacea-i.com to facilitate translation of these findings into clinical practice.

## Data Availability Statement

The data that support the findings of this study are available from the corresponding author upon reasonable request, subject to ethical approval and institutional data sharing agreements. The clinical decision support tool is accessible at https://mohs.panacea-i.com.

---

**Supplementary Figure 1.**
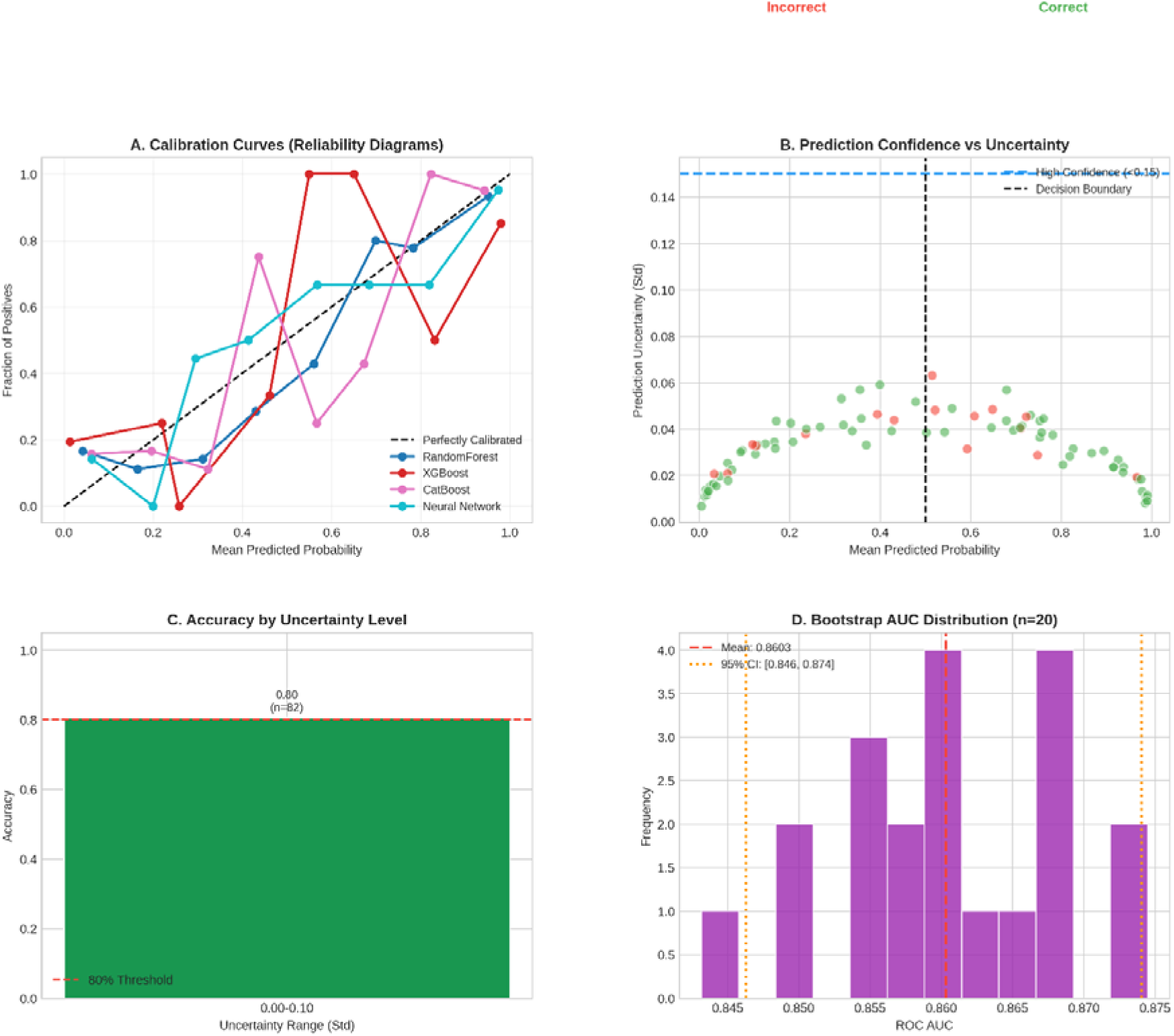
Model calibration and uncertainty analysis. (A) Calibration curves (reliability diagrams) for top models. Perfect calibration shown by diagonal. (B) Prediction confidence versus uncertainty scatter plot. Green points indicate correct predictions; red points indicate incorrect predictions. (C) Accuracy stratified by uncertainty level. (D) Bootstrap distribution of AUC values (n=20) with 95% confidence interval.

**Supplementary Table 1.**
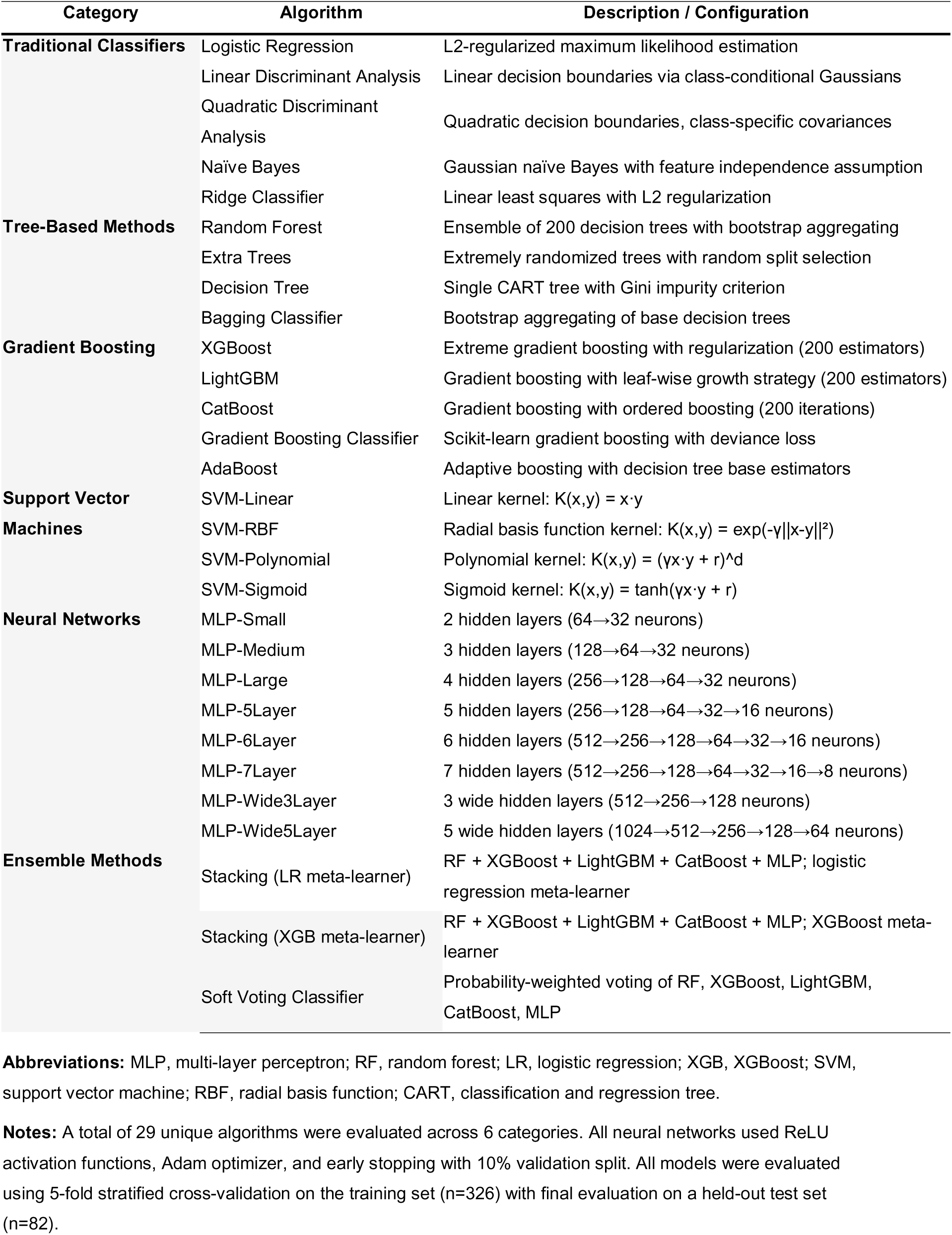
Machine Learning Algorithms Evaluated for Predicting ≥13 Sections.

